# Heart failure, female sex and atrial fibrillation are the main drivers of human atrial cardiomyopathy: results from the CATCH ME consortium

**DOI:** 10.1101/2023.03.23.23287667

**Authors:** J. Winters, A. Isaacs, S. Zeemering, M. Kawczynski, B. Maesen, J. Maessen, E. Bidar, B. Boukens, B. Hermans, A van Hunnik, B. Casadei, L. Fabritz, W. Chua, L.C. Sommerfeld, E. Guasch, L. Mont, M. Batlle, S. Hatem, P. Kirchhof, R. Wakili, M.F. Sinner, S. Kääb, M. Stoll, A. Goette, S. Verheule, U. Schotten

## Abstract

**Background:** Atrial cardiomyopathy (AtCM) is emerging as an independent prognostic factor in cardiovascular disease. Fibrotic remodeling, cardiomyocyte hypertrophy, and capillary density are histological hallmarks of atCM. However, the contribution of various etiological factors and atrial fibrillation (AF) to the development of differential atCM phenotypes has not been robustly quantified. We aimed to evaluate the association between histological features of atCM and the clinical phenotype.

**Methods:** We examined left (LA, n=95) and right (RA, n=76) atrial appendages sampled from a European cohort of patients undergoing cardiac surgery. Quantification of histological atCM features was performed using the JavaCyte algorithm, following staining with agglutinin (WGA), CD31 and vimentin. The contributions of AF, heart failure (HF), sex and age to histological characteristics were determined in a multivariate model. K-means clustering of 6 histological features was performed to identify different types of atCM.

**Results:** In both LA and RA, persistent AF was associated with increased endomysial fibrosis (LA:+1.07±0.41µm,p=0.01; RA:+0.89±0.43µm,p=0.032), whereas total extracellular matrix (ECM) content was unchanged in AF. Men had larger cardiomyocytes (LA:+1.87±0.72μm,p=0.012), while women had a higher degree of endomysial fibrosis (LA:+0.99±0.51µm,p=0.048). Heart failure patients showed more endomysial fibrosis (LA:+1.79±0.41µm,p<0.001) and ECM content (LA:+2.93±1.15%, p=0.014), and a higher capillary density (LA:+0.14±0.06,p=0.032) and size (LA:+0.48±0.23µm,p=0.041; RA:+0.31±0.16µm,p=0.047). Clustering of samples based on structural features identified 2 distinct atCM phenotypes; one characterized by enhanced endomysial fibrosis (LA:+3.35µm,p<0.001; RA:+1.88μm,p<0.001), ECM content (LA:+5.68%,p<0.001; RA:+7.78%,p<0.001), and a higher fibroblast density (LA:+4.79%,p<0,001) and one characterized by cardiomyocyte hypertrophy (LA:+1.20µm,p=0.009; RA:+2.95µm, p<0.001). Patients with fibrotic atCM were more often female (LA:OR=1.31,p=0.003; RA:OR=1.55,p=0.003), had more often persistent AF (LA:OR=1.23,p=0.031) or heart failure (LA:OR=1.62,p<0.001) whereas hypertrophic features were more common in men (LA:OR=1.31,p=0.031; RA:OR= 1.55,p=0.003).

**Conclusions:** AtCM phenotypes vary with patient characteristics. Fibrotic atCM is associated with female sex, persistent AF and heart failure, while hypertrophic features are more common in men.

## Introduction

Many cardiovascular diseases substantially impact atrial function. Heart failure, hypertension, supraventricular tachycardias, valvular diseases and thyrotoxicosis can, over time, deteriorate the mechanical, electrical and endocrine function of the atria. In some patients, these mechanisms cause a progressive atrial cardiomyopathy (atCM) which – in the consensus paper of Goette et al. in 2016 – was defined as “any complex of structural, architectural, contractile or electrophysiological changes affecting the atria with the potential to produce clinically relevant manifestations” (1). Importantly, atCM has a substantial impact on cardiac performance, and also on the occurrence of atrial fibrillation (AF) and stroke (2-4). Once AF occurs, the arrhythmia itself accelerates the progression of atCM (1, 5). AtCM mechanisms vary between individual patients and over time (6), likely contributing to the limited efficacy of rhythm control therapies for AF (7, 8).

One of the key features of atCM is enhanced fibrosis, as extensively reported in multiple animal models (9-17) and various clinical settings, including left/right ventricular dysfunction (18), AF (19, 20) or heart failure (21, 22). We recently reported that endomysial fibrosis, defined as extracellular matrix deposition between myocytes within the muscle bundles, rather than overall connective tissue content, is the main determinant of conduction disturbances in human AF (23). Fibroblasts are not only responsible for extracellular matrix formation, but they may also interact electrically with cardiomyocytes, thereby affecting conduction (24, 25). Increased cardiomyocyte size relates to increased wall thickness and chamber dilatation, both of which can alter electrical propagation (26-28). Finally, capillary rarefaction may be a marker of prolonged myocardial stress, as observed in patients with heart failure with preserved ejection fraction (HFpEF), hypertensive heart disease and AF (29, 30).

Although a first classification of atCM was proposed in the consensus paper from 2016 (31), a detailed investigation of the relationship between histological features of atCM and clinical traits is still missing. Here, we sought to comprehensively appraise histological changes in both right and left atrial samples in a large European cohort of patients undergoing cardiac surgery for a variety of indications. We aimed to systematically quantify the type of atrial fibrosis, fibroblast density, myocyte size, capillary density and capillary size using a previously validated, semi-automated quantification method developed for high throughput histological studies (32). Additionally, we aimed to cluster left and right atrial samples based on their histological features and to identify the clinical traits associated with these clusters.

## Materials and methods

### Patient selection and tissue sampling

Human left and/or right atrial appendage samples were collected from the CATCH-ME (www.catch-me.info) atrial tissue biobank of 227 patients undergoing coronary artery bypass grafting (CABG), valvular surgery or heart transplant surgery (HTX). An RNA sequencing data set of this study population was recently published (33). A total of 170 tissue samples (n=95 LA, n=76 RA, 20 of which were paired) were of sufficient quality and included in the analyses. The study was performed in accordance with relevant guidelines and was approved by the local ethical committees of the sampling centers.

### Triple staining and microscopy

Snap frozen tissue samples were cryo-sectioned (6μm) and stained with Wheat Germ Agglutinin (WGA), anti-CD31 and anti-vimentin, as previously described (32). Acetone-based antigen-fixation (10 min) was followed by blocking of non-specific binding sites with blocking solution (2m/v% fraction V BSA, 0.3M glycine) for 60 minutes. Rabbit monoclonal anti-vimentin (1/150, Abcam (ab92547), Cambridge) and mouse monoclonal anti-CD31 (1/50, Abcam (ab9498), Cambridge) were added overnight at room temperature to visualize fibroblasts and endothelial cells. After threefold washing with 1xPBS, a mixture of Alexa 594-conjugated WGA (1/200, ThermoFisher (W11262), Netherlands), goat anti-mouse 488 (1/200, ThermoFisher (A-10680), Netherlands) and goat anti-rabbit 405 (1/200, ThermoFisher (35550), Netherlands) was applied for 120 minutes. Prolong Gold Antifade mounting medium (ThermoFisher (P10144), Netherlands) was added when coverslips were mounted. Sections were imaged using a Leica microscope (DM4B) and camera (MC170HD). Only sections with transversely cut cardiomyocytes were selected and photographed for further analysis.

### Automated visualization and quantification of histological atCM features

Standardized, automated analysis of structural tissue characteristics was performed using the recently developed and validated custom-built ImageJ-plugin JavaCyte (32). An average of 3 images was investigated from 3 individual tissue slices per patient, totaling 547 WGA/CD31/vimentin positive images. Cardiomyocytes were detected by segmentation of objects based on the detection of local fluorescence minima. WGA images were thresholded applying Phansalkar’s algorithm for adaptive local thresholding (Figure 1). To quantify the overall connective tissue content, the fraction of extracellular matrix (total ECM) was calculated as the percentage of WGA-positive pixels. The distance between neighboring cardiomyocytes was quantified as a measure for endomysial fibrosis, as described previously (32). CD31 and vimentin-positive images were thresholded applying Landini’s HSB-based color threshold. Capillary density (capillaries per cardiomyocyte) and capillary minimal Feret diameter were obtained from thresholded CD31 images. The percentage of vimentin-signal was obtained from thresholded vimentin-positive images, after correction for CD31-vimentin double-positive pixels to rule out cross-reactivity with endothelial cells.

**Figure 1:**
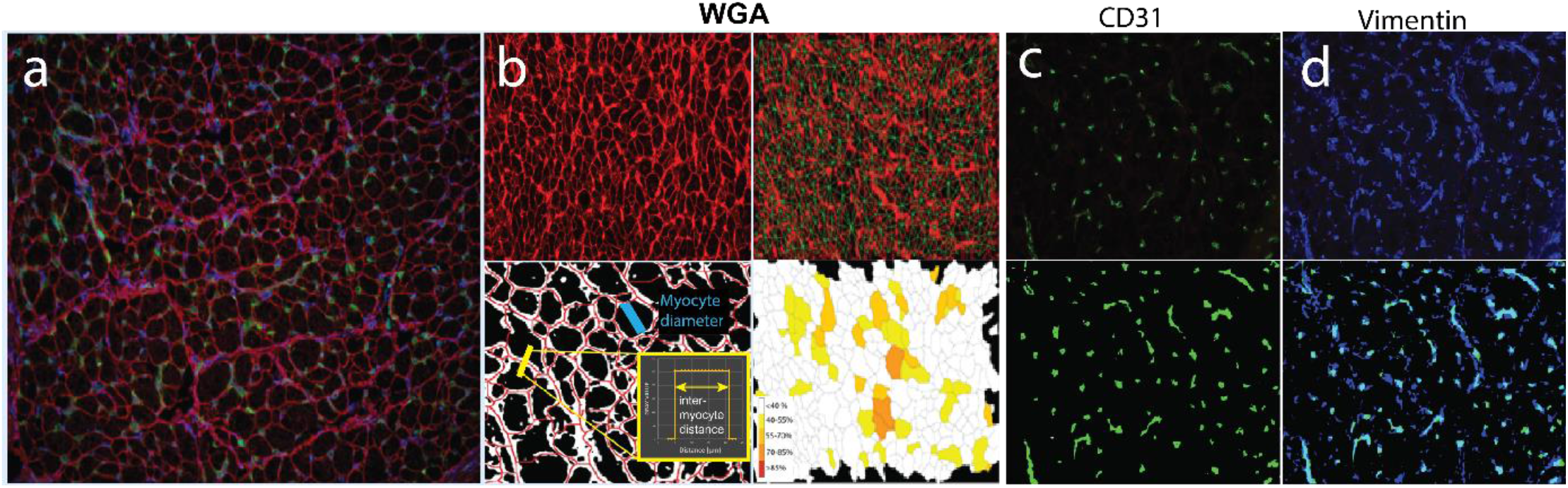
Methodology of immunohistochemical staining and the JavaCyte algorithm for analysis of histological parameters. a) Samples are triple-stained with WGA, CD31 and vimentin. b) After thresholding of the WGA image, the fraction of WGA-positive pixels is a measure for total ECM. Cell to cell distances were calculated as a measure of endomysial fibrosis. A control image is generated visualizing the measured distances between cells. Cardiomyocyte size is measured as the shortest diameter of the cells. Finally, a heatmap is generated with a color scale indicating the density of endomysial fibrosis surrounding each cardiomyocyte. Cluster analysis is performed to detect clusters of endomysial fibrosis. c) CD31 images are thresholded to count capillaries and normalize for cardiomyocyte count in the corresponding WGA image. Minimal Feret diameter of each capillary is measured. d) Vimentin images are thresholded to calculate the fibroblast to cardiomyocyte ratio.

### Statistical analysis: Association between clinical traits and histological features

The open access statistical software R (version R4.0.1) was used for all analyses (34). Normality of continuous traits was assessed using the Wilks-Shapiro test. Due to non-normality of some traits, differences between left and right atria were assessed using the Mann-Whitney U-test, while Fisher’s exact test was used to compare dichotomous traits.

To study differences in atrial histology between LA and RA samples, all samples (n=171) were combined for analysis using a multivariate model including the covariate ‘tissue location’. Additionally, the interaction between heart failure and tissue location (LA/RA) was studied. Next, multivariate regression models were constructed to study the association between histological features and clinical features, stratified by side, using the R package ‘lme4’. Age, sex, rhythm history and heart failure were explicitly modeled based on a-priori knowledge from available literature and univariate associations with histological traits. Dimensionality of the clinical database was reduced by transformation of 13 clinical traits into four principal components (PCs) which were included in the multivariate models. These traits included: weight, height, indication for CABG surgery, prior myocardial infarction, indication for aortic and mitral valve surgery, tricuspid valve insufficiency, thyroid dysfunction, hypertension, chronic kidney failure, diabetes and history of stroke or TIA. The ‘prcomp package’(35) was used to calculate these PCs. The proportion of variance accounted for by the first four PCs in the LA and RA samples was 55.8% and 56.6%, respectively. Finally, per atrium, samples were clustered based on histological features into two groups applying unsupervised k-means clustering, using the standard R-function ‘kmeans’. Associations between clinical characteristics and the identified clusters of histological features was studied applying logistic regression, adjusted for rhythm history, heart failure, sex and age.

## Results

### Tissue sample selection

A total of 260 samples from 227 patients were collected from the CATCH-ME biobank. Dropout occurred due to low tissue quantity (inability to perform cryosectioning, n=66), poor tissue quality detected by microscopy (n=19) or failure to pass built-in quality control from JavaCyte (n=4, Figure 2). A total of 171 samples (n=95 LA, n=76 RA) were of sufficient quality for analysis. Clinical and demographic information of the included patients is summarized in Table 1.

**Table 1:** Patient characteristics of donors of selected LA and RA samples. Differences were tested with Mann-Whitney U-tests for continuous variables and Fisher’s exact tests for dichotomous variables. NoAF: No history of AF, pAF: paroxysmal AF, persAF: persistent/permanent AF; AV/MV aortic/mitral valve; PVI: concomitant pulmonary vein isolation; NYHA: New Yeark Heart Association index; HTX: Heart Transplant Xenograft; CAD: Coronary artery disease; CABG: Coronary artery bypass graft; TIA: Transient ischemic attack. Values are presented as (mean ± SD) for continuous traits or in absolute number and percentage for dichotomous traits.

|  | *LA (n=95)* | *RA (n=76)* | *p-value* |
| --- | --- | --- | --- |
| *Sex: F* | 28 (29.3%) | 15 (18.9%) | 0.129 |
| *Age* | 63.0±11.0 | 65.3±10.7 | 0.201 |
| *Weight (kg)* | 77.7±15.1 | 80.2±16.4 | 0.309 |
| *Height (cm)* | 173±9.8 | 174±10.3 | 0.548 |
| *BMI* | 26.0±4.0 | 26.6±4.3 | 0.681 |
| *Rhythm* |  |  | 0.190 |
| • *NoAF* | 40 (42.1%) | 43 (56.6%) |  |
| • *pAF* | 29 (30.5%) | 18 (23.7%) |  |
| • *persAF* | 26 (27.4%) | 15 (19.7%) |  |
| *PVI* | 20 (21.1%) | 6 (7.9%) | **0.027** |
| *Heart failure* | 39 (40.2%) | 26 (33.8%) | 0.431 |
| *NYHA class* | 2.3±1.1 | 2.4±1.1 | 0.208 |
| *HTX* | 32 (33.7%) | 17 (22.4%) | 0.168 |
| *CHA<sub>2</sub>DS<sub>2</sub>-VASc* | 2.8±1.4 | 3.0±1.5 | 0.911 |
| *CAD* | 40 (42.1%) | 45 (59.2%) | **0.031** |
| *CABG* | 20 (21.1%) | 31 (40.8%) | **0.010** |
| *Prior MI* | 22 (23.2%) | 14 (18.4%) | 0.464 |
| *Diabetes* | 18 (18.9%) | 11 (14.5%) | 0.409 |
| *Chronic kidney failure* | 8 (8.4%) | 5 (6.6%) | 0.991 |
| *Hypertension* | 64 (67.4%) | 56 (73.7%) | 0.613 |
| *Prior stroke* | 10 (10.5%) | 4 (5.3%) | 0.274 |
| *Prior TIA* | 3 (3.2%) | 8 (10.5%) | 0.064 |
| *Thyroid disease* | 15 (15.8%) | 6 (7.9%) | 0.207 |
| *MV stenosis* | 3 (3.3%) | 3 (4.1%) | 0.973 |
| *MV regurgitation* |  |  | 0.871 |
| • *Grade 1* | 18 (19.6%) | 16 (21.6%) |  |
| • *Grade 2* | 14 (15.2%) | 10 (13.5%) |  |
| • *Grade 3* | 11 (12.0%) | 9 (12.2%) |  |
| • *Grade 4* | 13 (14.1%) | 6 (8.1%) |  |
| *MV surgery* | 26 (27.4%) | 14 (18.4%) | 0.222 |
| *AV stenosis* | 16 (17.4%) | 20 (27.0%) | 0.061 |
| *AV regurgitation* | 22 (17.4%) | 21 (28.9%) | 0.206 |
| *AV surgery* | 19 (20.0%) | 21 (28.9%) | 0.212 |
| *TV regurgitation* | 30 (31.6%) | 18 (23.7%) | 0.191 |

**Figure 2:**
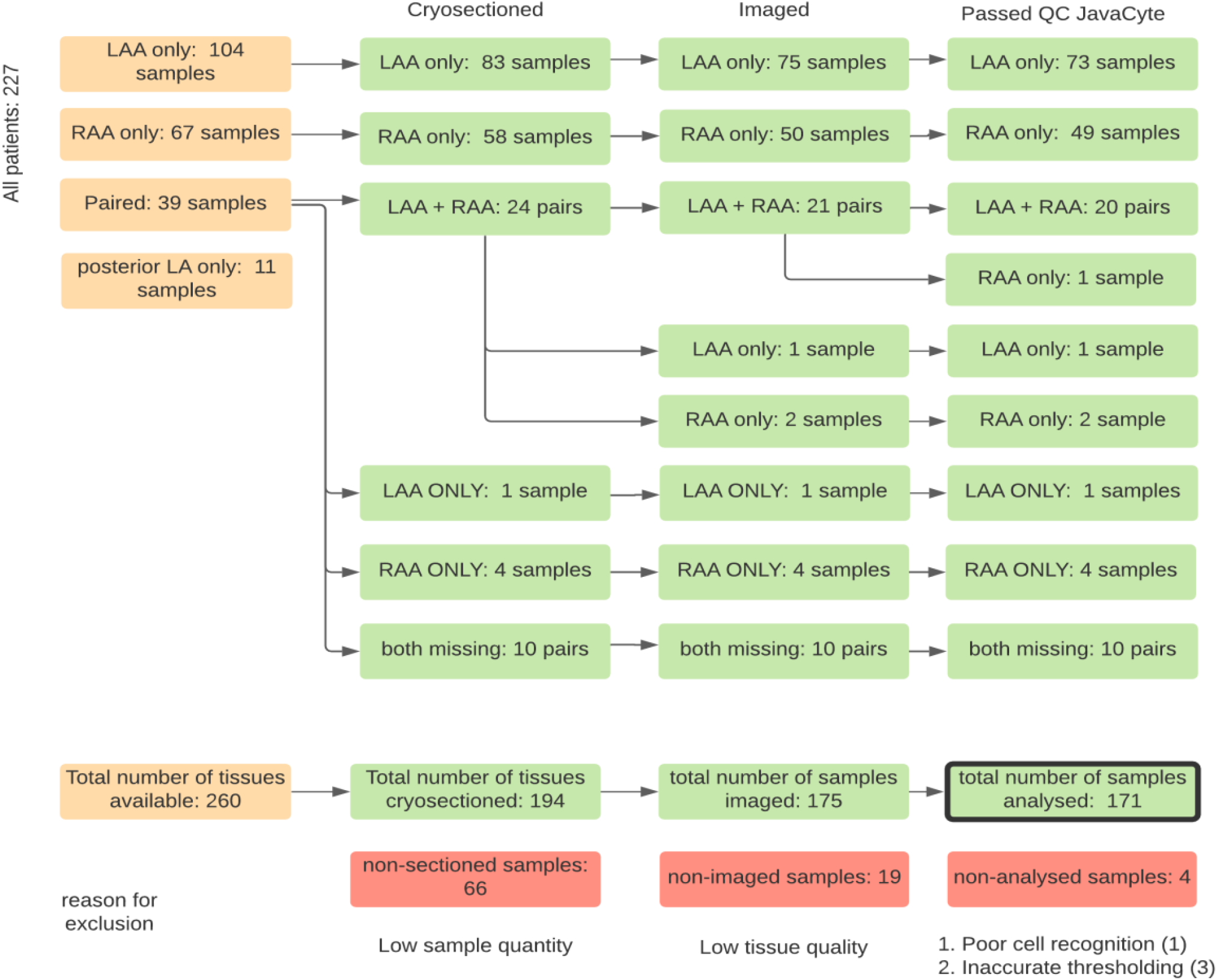
Flow chart of sample selection. 260 left or right atrial appendage (LAA/RAA) biopsies were collected from 227 patients. 66 samples were not cryosectioned because of low tissue quantity. 19 samples were excluded due to poor tissue quality observed during microscopy. An additional 4 samples were excluded as a result of failure to pass built-in quality control during analysis with JavaCyte. Reasons for exclusion were poor myocyte detection or poor thresholding of images.

### Right-left differences in atrial histology

Explorative univariate analysis showed that structural properties of LA and RA samples differed substantially. RA samples had more endomysial fibrosis (+1.28 μm, p<0.001) and a higher total extracellular matrix (ECM) content (+3.77%, p<0.001) than LA samples, but smaller cardiomyocytes (−0.69 μm, p=0.021). Non-significant trends towards less fibroblast-specific vimentin expression (−8.6%, p=0.092) and lower capillary density (−0.05 capillaries/myocyte, p=0.061) were observed in the RA. Capillary size did not differ between LA and RA samples (p=0.284). Multivariate regression analyses including the covariates age, sex, rhythm history, heart failure, four PCs and atrial biopsy location (after exclusion of paired samples) confirmed the RA/LA differences (Figure 3), including more endomysial fibrosis (+1.51 ±0.25 µm, p<0.001), a larger total ECM content (+6.15 ±1.14%, p<0.001), lower capillary density (−0.06±0.03 capillaries/myocyte, p=0.044), larger capillary size (+0.26±0.12µm, p=0.029), but smaller cardiomyocytes (−0.92±0.31 µm, p=0.003) in the RA. Moreover, we found a significant negative interaction (−2.52±0.73, p<0.001) between atrial side and heart failure status on endomysial fibrosis, indicating that heart failure is more strongly associated with endomysial fibrosis in LA than RA samples. Given the substantial difference in atrial histology between atria, further analyses were stratified by atrial side.

**Figure 3:**
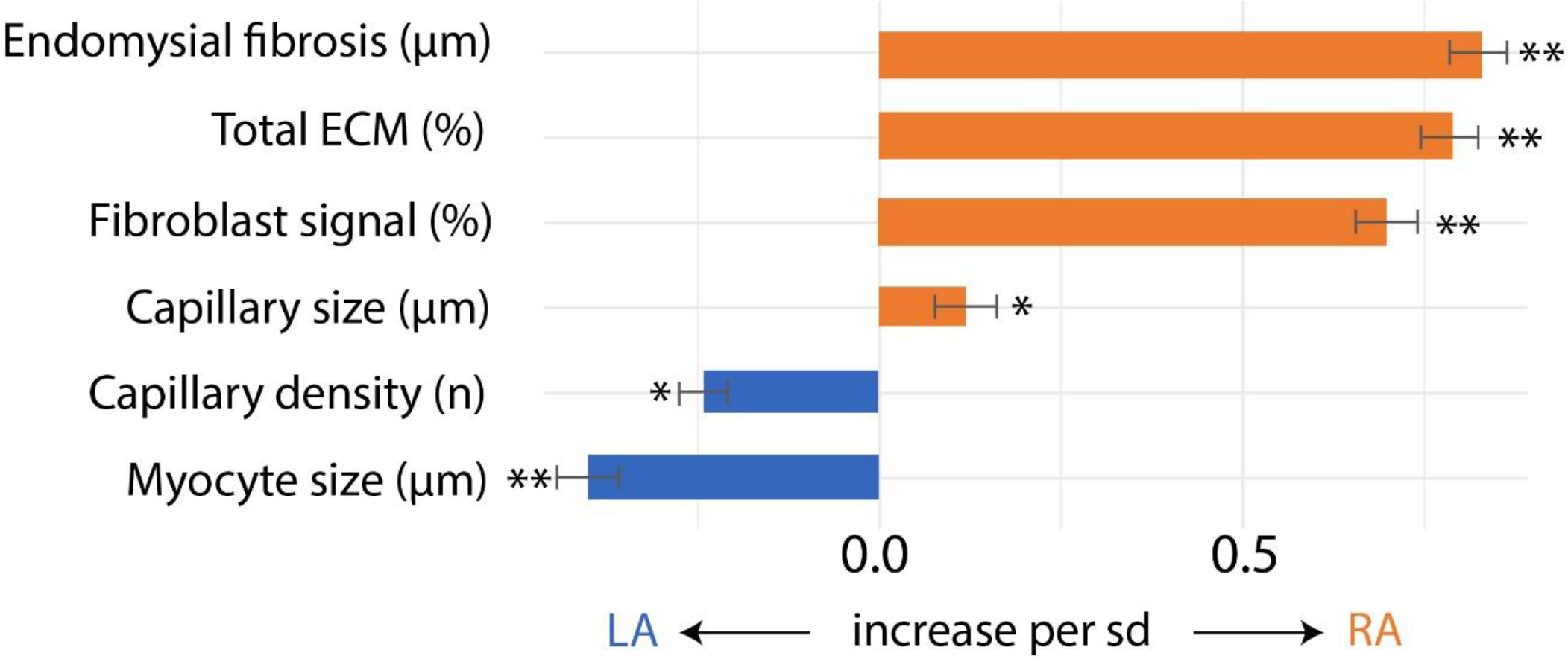
Differences in atrial histology between LA and RA samples. The effect of tissue sampling location (LA/RA) on histological parameters was tested in multivariate models including the covariates: rhythm, heart failure, age, sex, and 4 PCs. The effect size (β) was plotted relative to an increase in each histological parameter of 1 standard deviation. * p-value <0.05, ** p-value < 0.01

### Association between clinical traits and histological features of atCM

Explorative univariate analysis, stratified by atrial side, identified candidate traits associated with structural remodeling features (Supplemental Table 1). Multivariate regression was performed to study associations between histological features and clinical profiles (Figure 4, Supplemental Table 2). Rhythm history, heart failure, sex, and age were included as covariates in the multivariate analysis as they were univariately associated with clinical traits. Additionally, 4 principal components containing univariately associated clinical traits were also included. Overall, heart failure, rhythm history and female sex showed the strongest associations with atrial histology, with stronger associations in the LA than RA (Figure 4).

**Figure 4:**
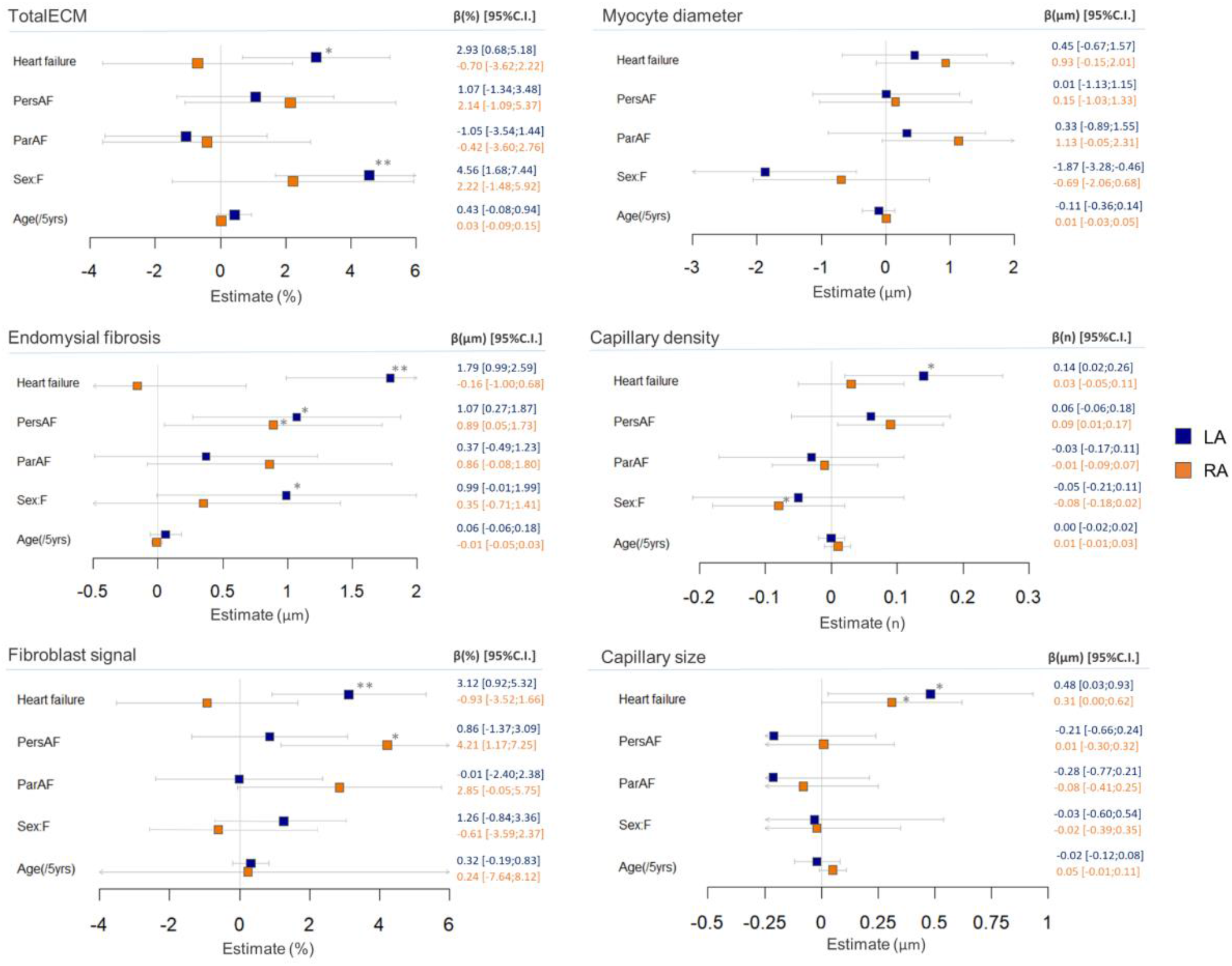
Results of multivariate models describing the association between clinical traits and histological features. Multivariate regression models were designed for each histological trait, stratified by atrial side. Sex, heart failure, rhythm history and age are modeled explicitly, supplemented by four principal components (PC) including additional clinical traits (not shown, not significant). Overall, heart failure, rhythm history and sex are the strongest drivers of structural remodeling in this dataset. Clinical traits have a slightly more pronounced effect on histology in the LA than RA. * p-value <0.05, ** p-value < 0.01

Female sex and heart failure were associated with higher ECM content in the LA (Figure 4). More endomysial fibrosis was observed in both atria of patients with persAF, in the LA of women, and in LA of patients with heart failure (Figure 4). More fibroblast-specific signal was detected in the RA of persAF patients and the LA of heart failure patients (Figure 4). Moreover, male patients had larger cardiomyocytes in the LA (Figure 4). Increased capillary density was observed in the LA of heart failure patients (Figure 4). An increased capillary size was observed in both atria of heart failure patients (Figure 4). Table 3 summarizes the numeric details of this analysis.

After exclusion of younger patients with end-stage heart failure receiving an HTX, age was associated with increased total ECM ratio (LA, p=0.022) as well as with enhanced endomysial fibrosis (LA: p=0.05 /RA: p=0.048). Capillary density was no longer increased in heart failure patients after exclusion on HTX-patients (Supplemental Tables 3 and 4).

A moderate (Pearson) correlation was observed between endomysial fibrosis and total ECM in the LA (r=0.68, p<0.001) and RA (r=0.60, p<0.001) samples (Figure 5).

**Figure 5:**
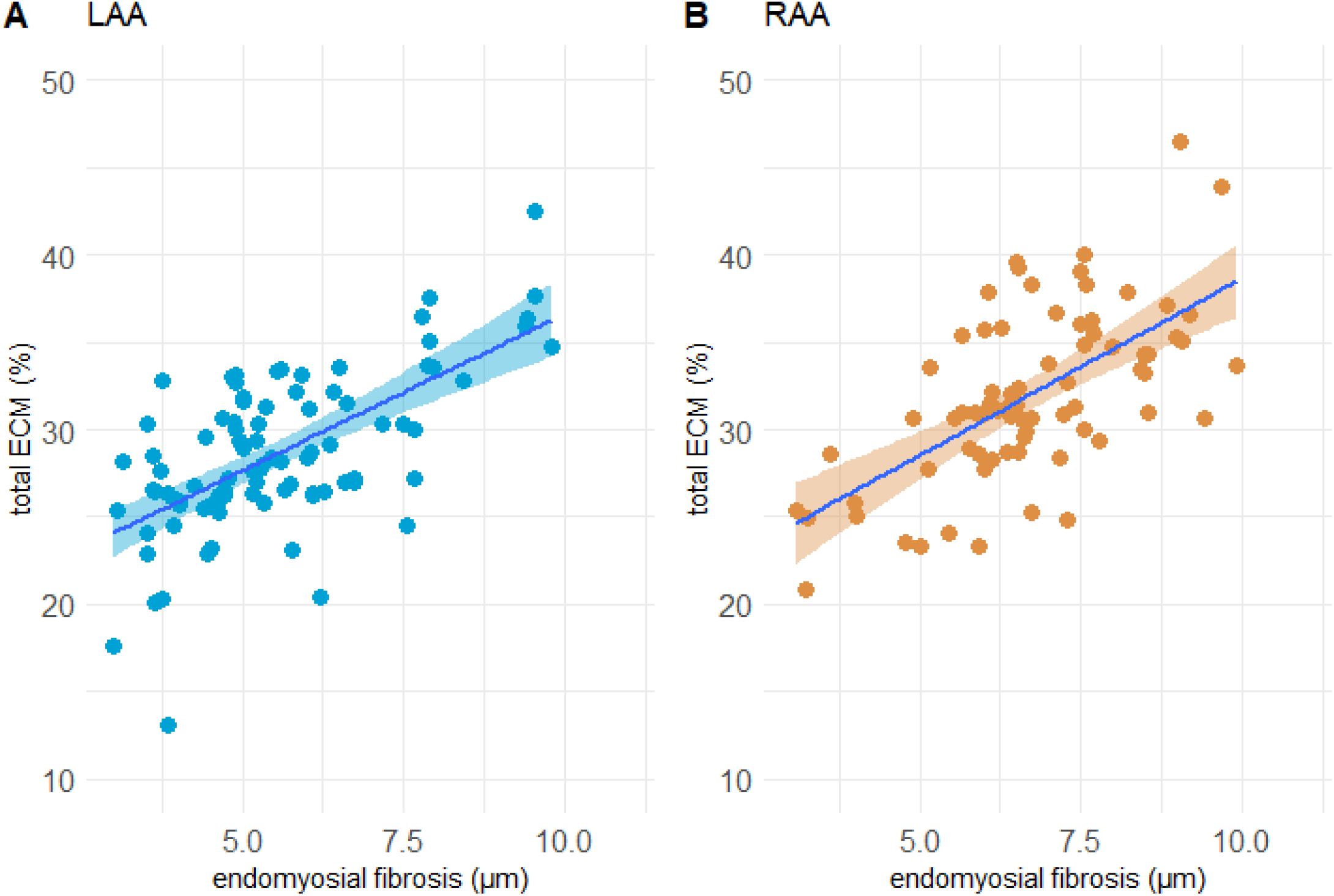
Correlation between total ECM and endomysial fibrosis in LA and RA samples. Endomysial fibrosis correlates moderately with total ECM (LAA: r=0.68, p<0.001), RAA: r=0.60, p<0.001). ECM: extracellular matrix content, LAA/RAA: left/right atrial appendage

### Clustering of left and right atrial samples based on structural features

Unsupervised k-means clustering was performed on left (n=91) and right (n=74) atrial samples to identify unique clusters based on fibrotic content (total ECM, endomysial fibrosis and fibroblast-specific vimentin expression), cardiomyocyte size, capillary density and size. Due to incomplete phenome data, 4 LA and 2 RA samples were excluded. Inspection of the WSS-plot and silhouette curve indicated 2 clusters per atrium (not shown). Structural properties of the samples within these clusters suggest that in each atrium, one cluster represents samples with more fibrosis (fibrotic atCM cluster), while the other cluster contains samples with larger cardiomyocytes (Figure 6, hypertrophic atCM cluster). LA samples grouped in the fibrotic cardiomyopathy cluster contained more total ECM content (p<0.001), endomysial fibrosis (p<0.001), and fibroblast-specific vimentin signal (p<0.001) and a higher capillary density (p<0.001) than samples in the hypertrophic cluster, which contained samples with larger cardiomyocytes (p=0.008). Capillary size did not differ. RA samples in the fibrotic cardiomyopathy cluster were characterized by more total ECM (p<0.001), more endomysial fibrosis (p<0.001), smaller cardiomyocytes (p<0.001) and a lower capillary density (p=0.012) than samples in the hypertrophic cluster. Fibroblast-specific vimentin signal and capillary size did not differ in the RA clusters (Figure 6).

**Figure 6:**
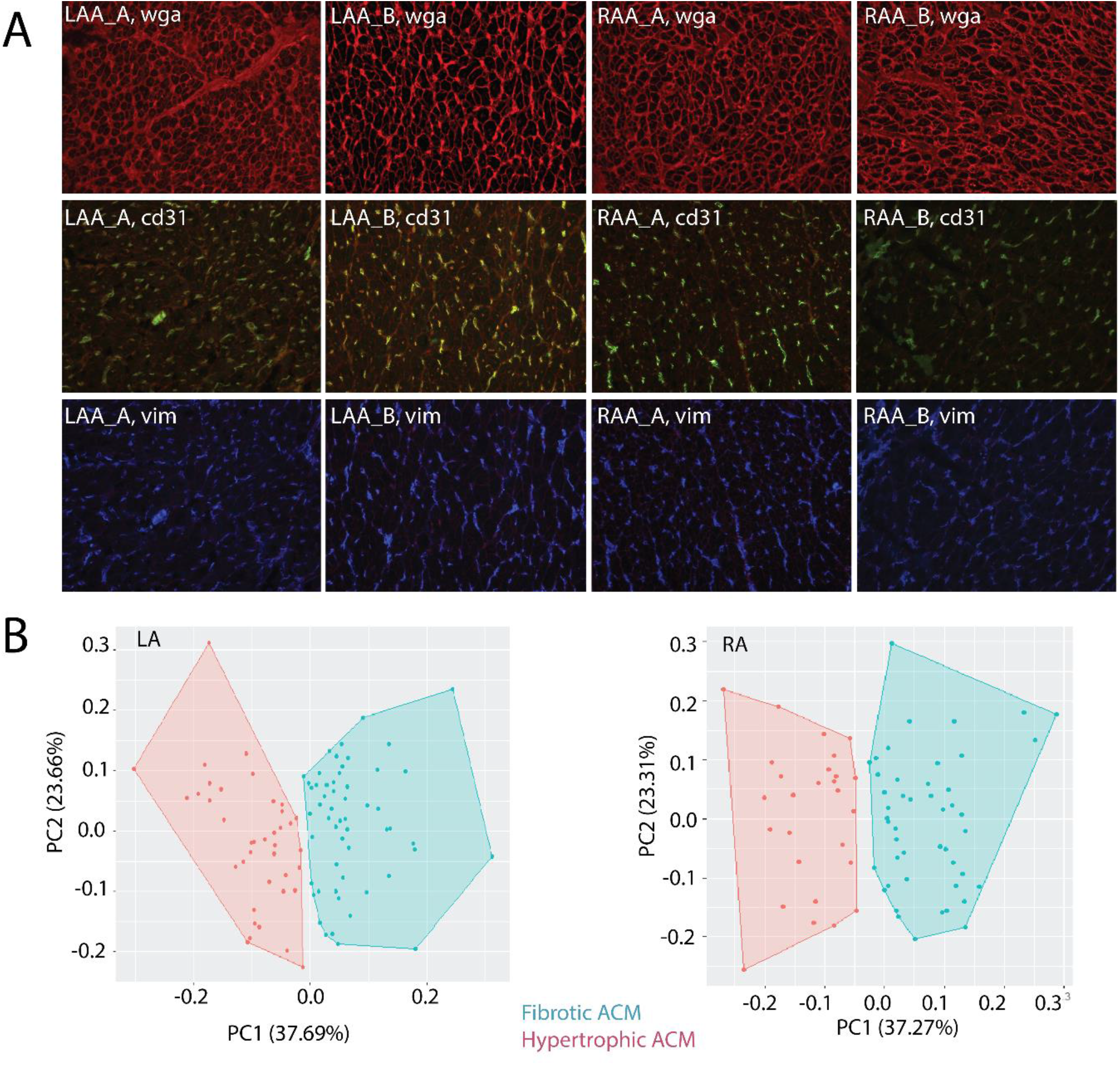
Clustering of LA and RA samples based on structural features. A) histological images of tissue features following staining with WGA, CD31 and vimentin. B) K-means clustering was applied to cluster LA and RA samples into 2 clusters per side. Total ECM and endomysial fibrosis were the main positive contributors to PC1, whereas myocyte diameter was the main negative contributor to PC1. Capillary density was the main contributor to PC2. WGA: wheat germ agglutinin; vim: vimentin; ECM: extracellular matrix content

Univariate analysis indicated that patients with fibrotic atCM in the LA more often had a history of AF (p=0.041) or heart failure (p<0.001). Heart failure with moderately or severely reduced left ventricular ejection fraction (HFrEF, p=0.003), in particular, was more common in this cluster (Supplemental Table 5). Moreover, the primary indication for surgery differed between patients showing fibrotic and hypertrophic atCM in the LA (p=0.007), with more coronary artery bypass grafting (CABG) or aortic valve surgery in patients with hypertrophic cardiomyopathy, but more HTX in patients with fibrotic cardiomyopathy (Supplementary Table 5). Patients with fibrotic remodeling in the RA were more often female (p=0.002) and shorter (p=0.042).

Multivariate logistic regression in LA samples revealed that, compared to hypertrophic atCM, patients with fibrotic atCM were more often female (OR: 1.31, SE: 0.09, p=0.003), and had a history of persistent AF (OR: 1.23, SE:0.09, p=0.031) or heart failure (OR: 1.62, SE:0.08, p<0.001). Patients with fibrotic atCM in the RA were also more frequently female (OR: 1.55, SE: 0.14, p=0.003), but did not differ in history of persistent AF (OR: 1.06, SE: 0.15, p=0.668) or heart failure (OR: 0.86, SE: 0.14, p=0.233). Patients with hypertrophic atCM in the RA were more often male (OR: 1.55, SE:0.14, p=0.003), (Figure 7).

**Figure 7:**
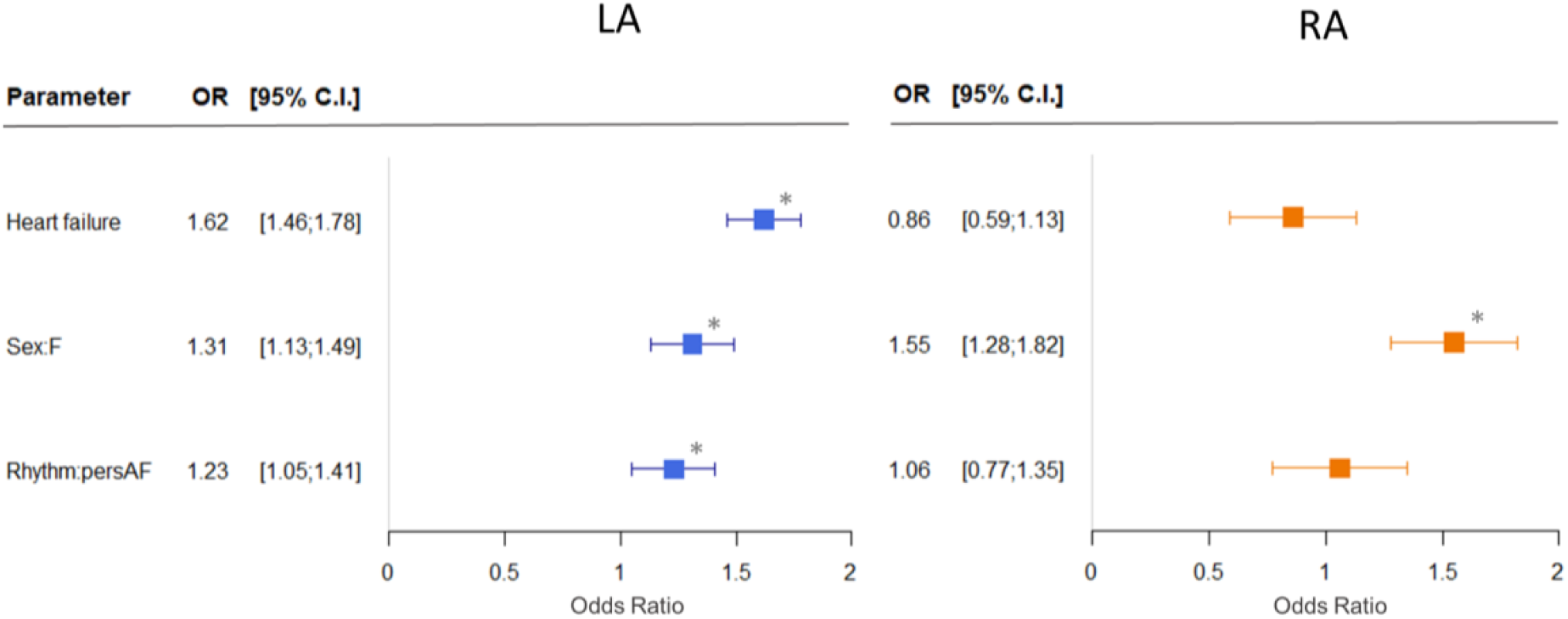
Distribution of clinical traits in fibrotic cardiomyopathy compared to hypertrophic cardiomyopathy. Patients with fibrotic cardiomyopathy in the LA were more often female, and more frequently had a history of persAF or heart failure. Fibrotic cardiomyopathy in the RA was more frequent in female patients. OR: odds ratio, C.I.: confidence interval.

## Discussion

The aim of this study was to better understand the relationship between the main AF-related comorbidities and clinical traits and histological features of atCM in right and left atrial tissue samples. We investigated various hallmarks of atCM in a large European cohort of patients with a variety of indications for cardiac surgery. We found that female sex, heart failure, and AF are the main drivers of fibrosis in human atria. Endomysial fibrosis, however, was independently associated with AF whereas over-all ECM content was not. Heart failure showed the strongest association with structural characteristics studied in this cohort, associated with higher total ECM content (LA), increased amounts of endomysial fibrosis (LA), higher capillary density (LA), higher cardiomyocyte size (LA/RA), and higher fibroblast density (LA). Contrary to earlier reports describing enlarged cardiomyocytes in dog models of CHF (20), we did not find an association between cardiomyocyte size and heart failure. We identified two distinct clusters of samples, one showing fibrotic cardiomyopathy and one demonstrating a hypertrophic phenotype. Fibrotic cardiomyopathy was associated with female sex, persistent AF and heart failure while hypertrophic cardiomyopathy more often occured in men and in patients undergoing coronary artery bypass graft or aortic valve surgery.

### Differences in myocardial structure in the LA versus RA

We observed more total ECM and endomysial fibrosis, lower capillary density, as well as larger capillaries and smaller cardiomyocytes, in RA samples compared to LA. In most published studies, histological assessment of the atrial myocardium is limited to either RA or LA. As a result, literature on direct right/left comparisons of atrial structural properties in patients is sparse. Our observation of a higher level of fibrosis in the RA is in line with an earlier report based on immunohistochemical observations (36). A post-mortem analysis by Platonov et al. extensively studied atrial wall samples from five locations in the right and left atrium and did not observe any difference in fibrotic content, capillary density or cardiomyocyte size between right and left atrial locations (37). A recent late gadolinium-enhancement cardiac magnetic resonance imaging study (LGE-MRI) showed similar fibrotic levels in right and left atria and a strong correlation between them (38). At this point, the mechanistic reason for the high fibrotic levels reported in RA samples reported here remains unknown.

### Correlation between endomysial fibrosis and total ECM content

Prior research by our group showed that endomysial fibrosis, but not total ECM content, correlates well with AF complexity, i.e. the degree of conduction block in fibrillating atria (23). In the present study, the correlation between total ECM and endomysial fibrosis was significant but only moderately strong. This suggests that, in order to assess the contribution of atrial fibrosis to the electrophysiological substrate for AF, determining endomysial fibrosis may be more relevant than total ECM content quantification. In addition, it is unknown whether LGE-MRI can accurately quantify endomysial fibrosis; it relies on regional differences in a shift in T1 time of the gadolinium-based contrast agent, which may only be sensitive enough to detect thick strands of connective tissue (39-41). To a large extent, those large strands of connective tissue form the physiological skeleton of the atrium and do not necessarily contribute to the electrophysiological substrate for AF. Indeed, the contribution of the diffuse and thin layers of endomyisal fibrosis to the LGE-MRI signal is likely to be minimal. This stresses the need for identifying other markers for endomysial fibrosis, such as non-invasive electrophysiological characteristics or circulating biomolecules.

### Heart failure and structural remodeling

Our data confirm previous observations that heart failure is a strong determinant of structural alterations in the atria with an increase in total ECM content, endomysial fibrosis, fibroblast-specific signal, and capillary density and size. Furthermore, these data suggest that heart failure-associated structural changes are more prominent in the LA than in the RA, most likely as a consequence of chronically increased atrial wall stress caused by pressure or volume overload (21), which, in most patients, is limited to the LA. This hypothesis is supported by the notion of localized conduction heterogeneities due to increased LA fibrosis in dogs or sheep with tachypacing-induced heart failure (17, 22). Additionally, we report more fibroblast-specific vimentin staining in LA samples of heart failure patients. Fibroblasts not only produce extracellular matrix (ECM) causing fibrosis, but can also couple with myocytes potentially promoting reentry and ectopic activity (25, 42-45).

We observed larger capillary density (LA) in heart failure patients. Pathological pressure or volume overload stretches cardiomyocytes, activating pro-hypertrophic and pro-angiogenic signaling proteins to cope with increased oxygen demands (46-48). Consequently, the observed capillary surplus in atrial myocardium of heart failure patients described here could be indicative of an angiogenic response. After exclusion of patients receiving a transplant, capillary density was not altered in heart failure patients. AF and HFpEF patients often show decreased myocardial capillary density (49, 50), resulting in a lower coronary flow reserve and a stiffer myocardium, which are important contributors to diastolic dysfunction (29, 51-54).

### AF and structural remodeling

Our data showed an independent association between AF and endomysial fibrosis in human LA and RA, while AF was not associated with total ECM, altered myocyte size, capillary density or fibroblast signal. Structural remodeling of atrial myocardium was assessed in animal models of AF by rapid atrial pacing (9-14) and of congestive heart failure by ventricular tachypacing (15-17, 55). Ventricular tachypacing models are characterized by rapid, pronounced development of atrial fibrosis, whereas rapid atrial pacing models initially show less pronounced fibrosis (17, 56, 57). Fibrosis in ventricular tachypacing models is characterized by thick, extended strands of collagen fibers occurring after an initial phase of cardiomyocyte apoptosis (17, 58), indicative of replacement fibrosis. In goats, after 6 months of rapid atrial pacing-induced AF, atrial endomysial fibrosis increases, whereas quantities of total ECM remain constant (9), a combination of findings resembling well the findings of the current study. Recently, a study of cardiac surgery patients reported no association between electrophysiological parameters and histological fibrosis markers (59). Electrophysiological measurements, however, were performed at low rates and a distinction between total and endomysial fibrosis was only made in a small percentage of patients. Maesen et al. demonstrated that among several structural alterations in the atria, endomysial fibrosis, but not total ECM, is the strongest determinant of the degree of conduction block and AF complexity (23).

From the association between atrial endomysial fibrosis and AF observed here, we cannot infer causality. Structural remodeling, and endomysial fibrosis in particular, was described as consequence of AF in experimental studies (9, 10, 20, 60). On the other hand, several studies highlighted the role of both longitudinal (61) and transverse electrical uncoupling resulting from endomysial fibrosis in discontinuous conduction, microreentry (9, 54, 62, 63), and local heterogeneities of conduction (9, 12, 17, 23, 64). Persistent AF may, therefore, be both a cause and a consequence of endomysial fibrosis.

### Aging and atrial structural remodeling

While Gramley et al. reported an increase in replacement fibrosis with increasing age in RA samples from cardiac surgery patients (65, 66), an extensive study by Platonov et al. reported no correlation between age and fibrosis in post-mortem human atrial samples obtained from the pulmonary veins, posterior left atrial wall, terminal crest and Bachmann’s bundle (37). By contrast, both studies reported a correlation between fibrosis and AF persistence. In their investigations, neither Gramley nor Platonov measured endomysial fibrosis, but quantified total connective tissue content. In the present study, an association between age and endomysial fibrosis was observed in heart failure patients who did not receive a transplant. Electrical uncoupling of cardiomyocytes (67), enhanced anisotropy of conduction velocity (62), and increased fibrosis (68, 69) were observed as a result of ageing in dogs. Spach et al. found local slowing of transverse, but not longitudinal, conduction in atrial muscle bundles of elderly patients in whom micro-fibrosis was more pronounced compared to younger patients (70, 71). Our, and the referenced experimental data, suggest that endomysial fibrosis might contribute to the association between AF and age to some degree. In the present study, however, the relative contribution of age was quantitatively limited. A 5-year age difference, causing a 1.5-fold increase in AF prevalence in the general population (72), was associated with an 0.06 µm increase in endomysial fibrosis in our study (comparable to 3.3% or 5.6% of the effect of heart failure or persistent AF).

### Sex-difference in atrial fibrosis

In AF patients, female sex is associated with a higher risk of stroke (73), higher recurrence rates following ablation (74), and increased mortality (75), but the underlying pathophysiological mechanisms remain elusive. Several LGE-MRI studies demonstrated an independent association of female sex and atrial fibrosis (76-78) but it is unclear whether LGE-MRI can distinguish between different forms of fibrosis. There is only limited light microscopy data available on sex-differences in fibrosis in atrial tissue. In patients undergoing mitral valve surgery, increased fibrosis in the pulmonary vein sleeves was observed in female patients with AF compared to those without AF while such a difference was not found in males (79). Our study is the first to demonstrate a clear association between female sex and both LA total ECM and endomysial fibrosis. Interestingly, the effect size was comparable to that of persistent AF or heart failure.

### Fibrotic versus hypertrophic cardiomyopathy

Clustering methods identified fibrotic and hypertrophic atCM as distinct types of atrial structural remodeling in our cohort. Patients with fibrotic atCM were more often women and were more likely to have persistent AF or heart failure, while patients with hypertrophic atCM were more often men or patients undergoing coronary artery bypass graft or aortic valve surgery. This result can be seen as a first step towards the development of a classification of atCM based on histological features and clinical traits associated with them. To a certain degree, our results support aspects of the EHRA’s classification proposed in the consensus paper in 2016 (1). Our data confirm the association between fibrotic alterations and AF or heart failure, while other aspects identified by our study, such as sex-differences in endomysial fibrosis and cardiomyocyte size, were not included in the EHRA classification (1).

Our study also stresses the need for high throughput, standardized, automated histological analysis of RA and LA samples. Given the diversity of comorbidities that impact atCM, only dedicated, large-scale, multi-centre studies using validated high-throughput histological quantification methods allow full validation or refinement of the EHRA classification of atCM and the assessment of its value in guiding clinical management of AF. The long-term vision remains that a more precise, ideally mechanistic, classification of atrial pathologies may contribute to the development of individualized therapeutic approaches to AF, potentially improving outcomes.

### Limitations

Data regarding pre-operative medication or nicotine and alcohol consumption were, unfortunately, not complete and confounding of our data by these factors cannot be ruled out. Also, we did not assess the role of fat tissue, inflammatory changes or endothelial remodeling. Additionally, the arrhythmogenic substrate often originates in the atrial sleeves of the pulmonary veins and the posterior wall of the left atrium, but tissue from these regions were not available. Histological properties of atrial appendages might not be fully representative for structural changes in other areas of the atria.

## Data Availability

The first author has full access to all the data in the study and verified the integrity of the primary data and any analyses performed.

---

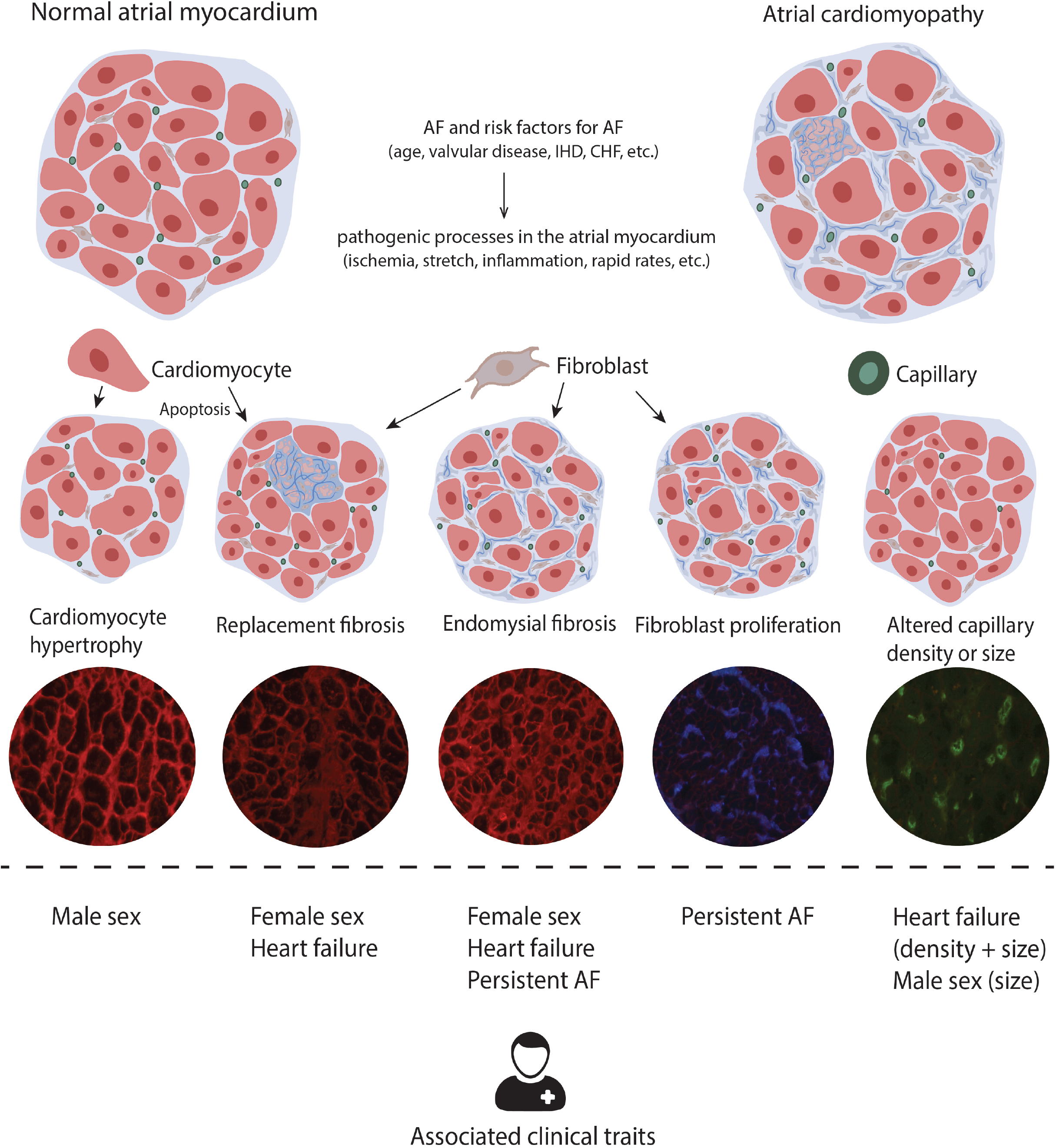

